# Excess mortality in mental health service users during the COVID-19 pandemic described by ethnic group: South London and Maudsley data.

**DOI:** 10.1101/2020.07.13.20152710

**Authors:** Robert Stewart, Matthew Broadbent, Jayati Das-Munshi

## Abstract

The COVID-19 pandemic in the UK was accompanied by excess all-cause mortality at a national level, only part of which was accounted for by known infections. Excess mortality has previously been described in people who had received care from the South London and Maudsley NHS Foundation Trust (SLaM), a large mental health service provider for 1.2m residents in south London. SLaM’s Clinical Record Interactive Search (CRIS) data resource receives 24-hourly updates from its full electronic health record, including regularly sourced national mortality on all past and present SLaM service users. SLaM’s urban catchment has high levels of deprivation and is ethnically diverse, so the objective of the descriptive analyses reported in this manuscript was to compare mortality in SLaM service users from 16^th^ March to 15^th^ May 2020 to that for the same period in 2019 within specific ethnic groups: i) White British, ii) Other White, iii) Black African/Caribbean, iv) South Asian, v) Other, and vi) missing/not stated. For Black African/Caribbean patients (the largest minority ethnic group) this ratio was 3.33, compared to 2.47 for White British patients. Considering premature mortality (restricting to deaths below age 70), these ratios were 2.74 and 1.96 respectively. Ratios were also high for those from Other ethnic groups (2.63 for all mortality, 3.07 for premature mortality).

## Background

The COVID-19 pandemic is likely to have had a profound impact on people using mental healthcare because of the heightened vulnerability of its patient populations (e.g. through cardiovascular and respiratory disorders), already-reduced life-expectancies (1), and frequently described problems accessing healthcare (2;3). There is therefore a pressing need for information in the public domain (4). This manuscript is one of what we envisage will be a series of open-access reports to be generated, covering different elements of mental health service activity and outcomes from a large mental healthcare provider in south London, UK. The reports take advantage of relatively rapid access to source data and will be kept updated for as long as a single site’s experiences are deemed useful.

We have previously described a 2.4-fold excess mortality in past and present mental healthcare services users during the period of the pandemic experienced in London from 16^th^ March to 15^th^ May 2020 compared to mortality for the same 2-month period in 2019 (5). Because of well-publicised international concerns, we further investigated differences in excess mortality between ethnic groups.

## Methods

The Biomedical Research Centre (BRC) Case Register at the South London and Maudsley NHS Foundation Trust (SLaM) has been described previously (6;7). SLaM serves a geographic catchment of four south London boroughs (Croydon, Lambeth, Lewisham, Southwark) with a population of around 1.2 million residents, and has used a fully electronic health record (EHR) across all its services since 2006. SLaM’s BRC Case Register was set up in 2008, providing researcher access to de-identified data from SLaM’s EHR via the Clinical Record Interactive Search (CRIS) platform and within a robust security model and governance framework (8). CRIS has been extensively developed over the last 10 years with a range of external data linkages and natural language processing resources (7). Of relevance to the work presented here, CRIS is updated from SLaM’s EHR every 24 hours and thus provides relatively ‘real-time’ data. Mortality in the complete EHR (i.e. all SLaM patients with records, past or present) is ascertained weekly through automated checks of National Health Service (NHS) numbers (a unique identifier used in all UK health services) against a national spine. CRIS has supported over 200 peer reviewed publications to date. CRIS has received approval as a data source for secondary analyses (Oxford Research Ethics Committee C, reference 18/SC/0372).

Mortality data were quantified for the two month period from 16^th^ March to 15^th^ May for the years 2020 and 2019. These were described according to recorded ethnicity, categorised into the following groups from the source code categories: i) White British; ii) Other White (Irish, Any other white background); iii) Black African/Caribbean (Caribbean, African, Any other Black background); iv) South Asian (Indian, Pakistani, Bangladeshi); v) Other (White and Black Caribbean, White and Black African, White and Asian, Any other mixed background, Any other Asian background, Chinese, Any other ethnic group); vi) Unknown (missing or recorded as not stated). Numbers of deaths in the comparison years were described both as numeric differences and ratios. ‘Premature’ deaths were then described and evaluated in the same way, where the age at death was less than 70 years. Ethnic group distributions were then described for the following samples: i) total deaths in 2020; ii) excess deaths in 2020 (i.e. deaths in 2020 minus those in 2019); iii) total premature deaths in 2020; iv) excess premature deaths in 2020.

It should be noted that the mortality data extraction was carried out at a later date (18^th^ June 2020) for this analysis than that for a previous report covering the same period (5); numbers of deaths may therefore differ due to updates on death registrations.

## Results

Data on numbers of deaths in 2020 in total and by ethnic group are displayed in Table 1. For the SLaM sample as a whole there were 968 more deaths between 16^th^ March and 15^th^ May 2020 than there were for the same period in 2019, and 215 more premature deaths. Taking into account the 61 days evaluated in both years, the total excess deaths in 2020 amounted to an additional 82 days’ deaths at 2019 levels, and the excess premature deaths in 2020 were equivalent to an additional 50 days at 2019 levels.

**Table 1:**
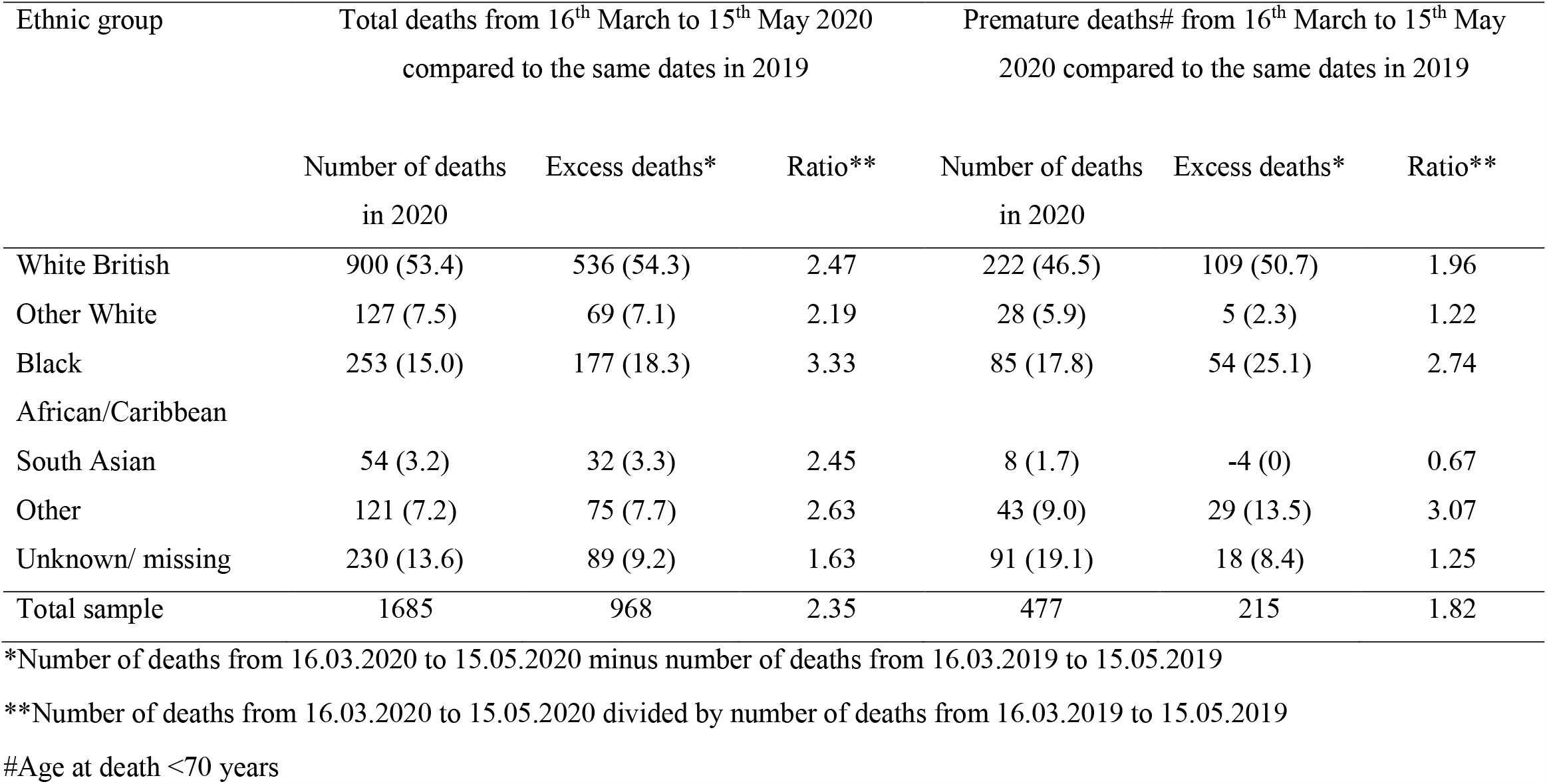
Comparisons between ethnic groups of lockdown-era deaths in 2020 compared to those seen in 2019 for previous/current SLaM service users

Ratios of 2020 to 2019 deaths were highest in the Black African/Caribbean group (3.33), followed by Other (2.63), White British (2.47), South Asian (2.45), Other White (2.19), and unknown (1.63) groups. For premature mortality, the ratios of 2020 to 2019 deaths were highest in the Other (3.07) group, followed by Black African/Caribbean (2.74), White British (1.96), Unknown (1.25) and Other White (1.22) groups, and the South Asian group had lower numbers of these deaths in 2020 than 2019. Ratios are compared graphically in Figure 1, and 2019/2020 daily mortality numbers in each group is displayed in Figures 2-7.

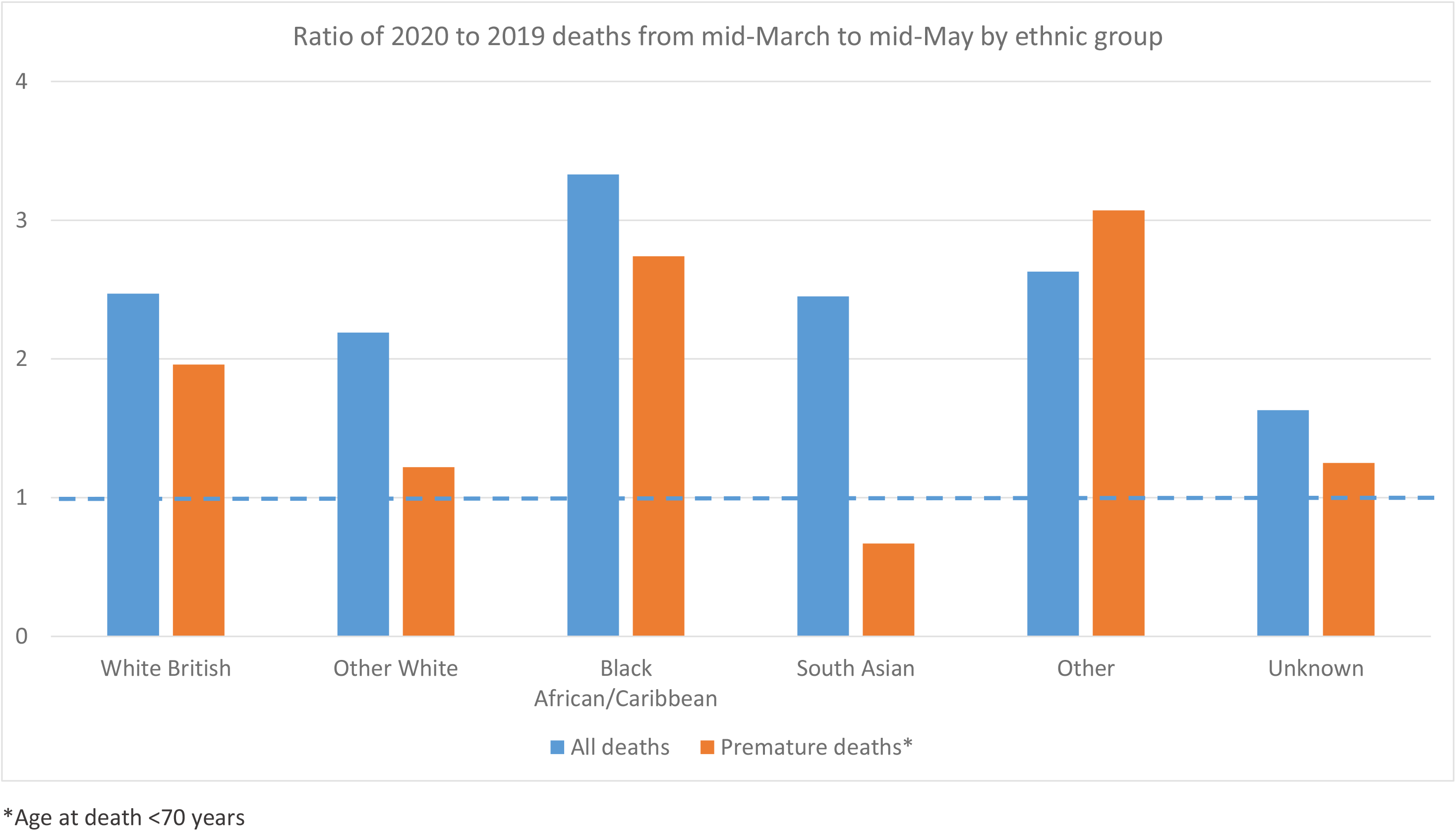

**Figure 2:**
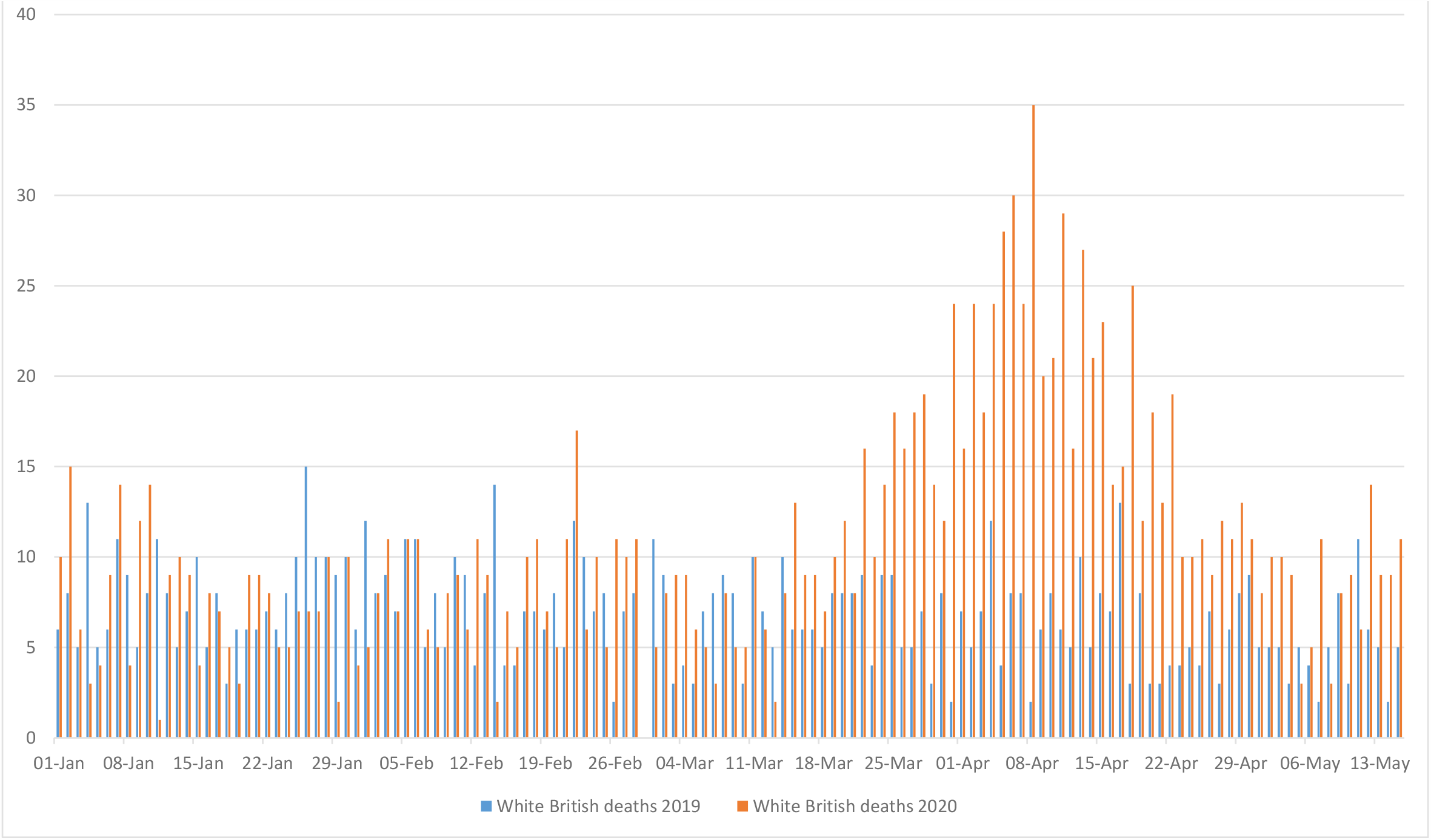
Daily mortality in mental health service users 2019 and 2020 -White British.

**Figure 3:**
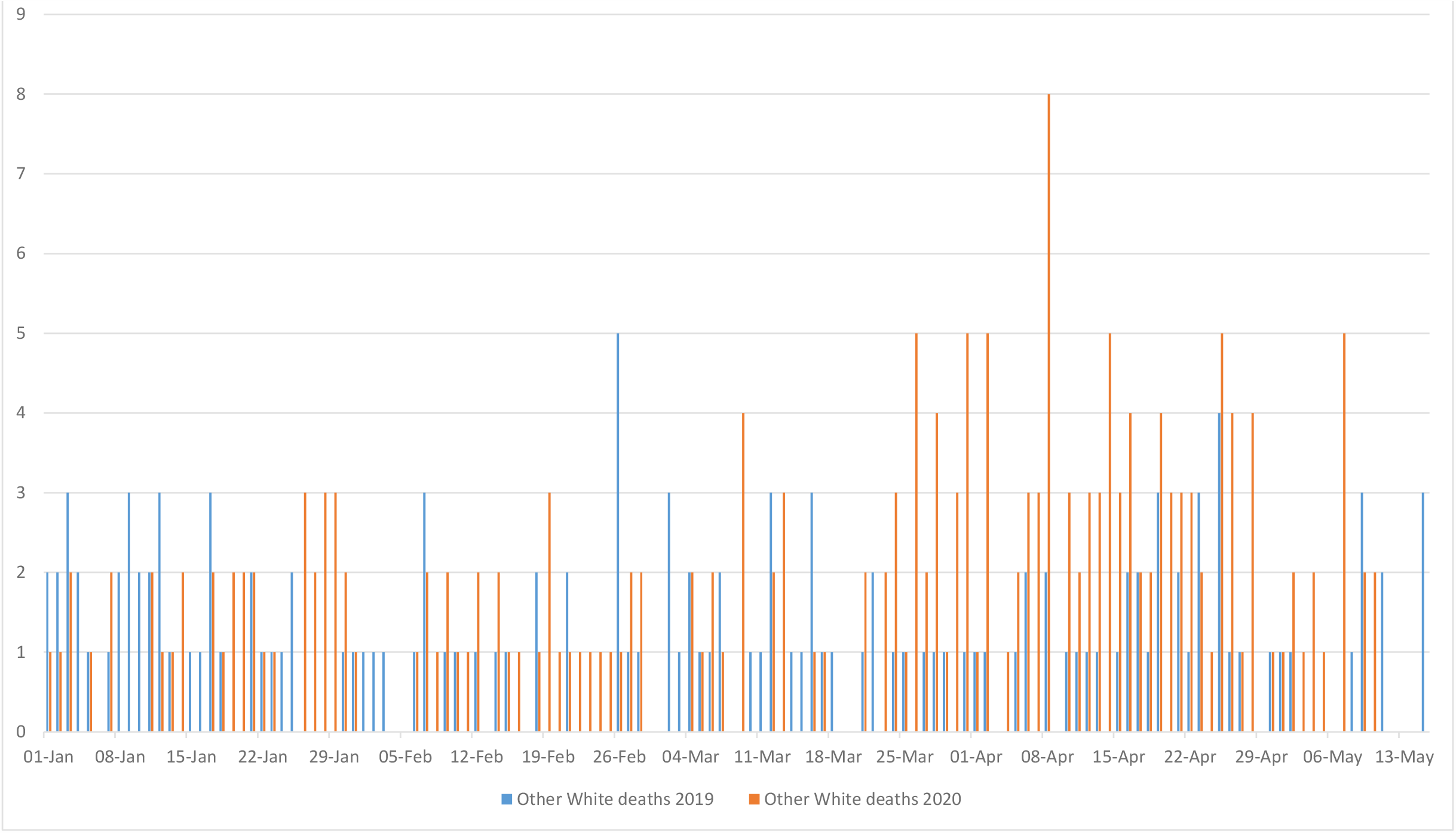
Daily mortality in mental health service users 2019 and 2020 -Other White.

**Figure 4:**
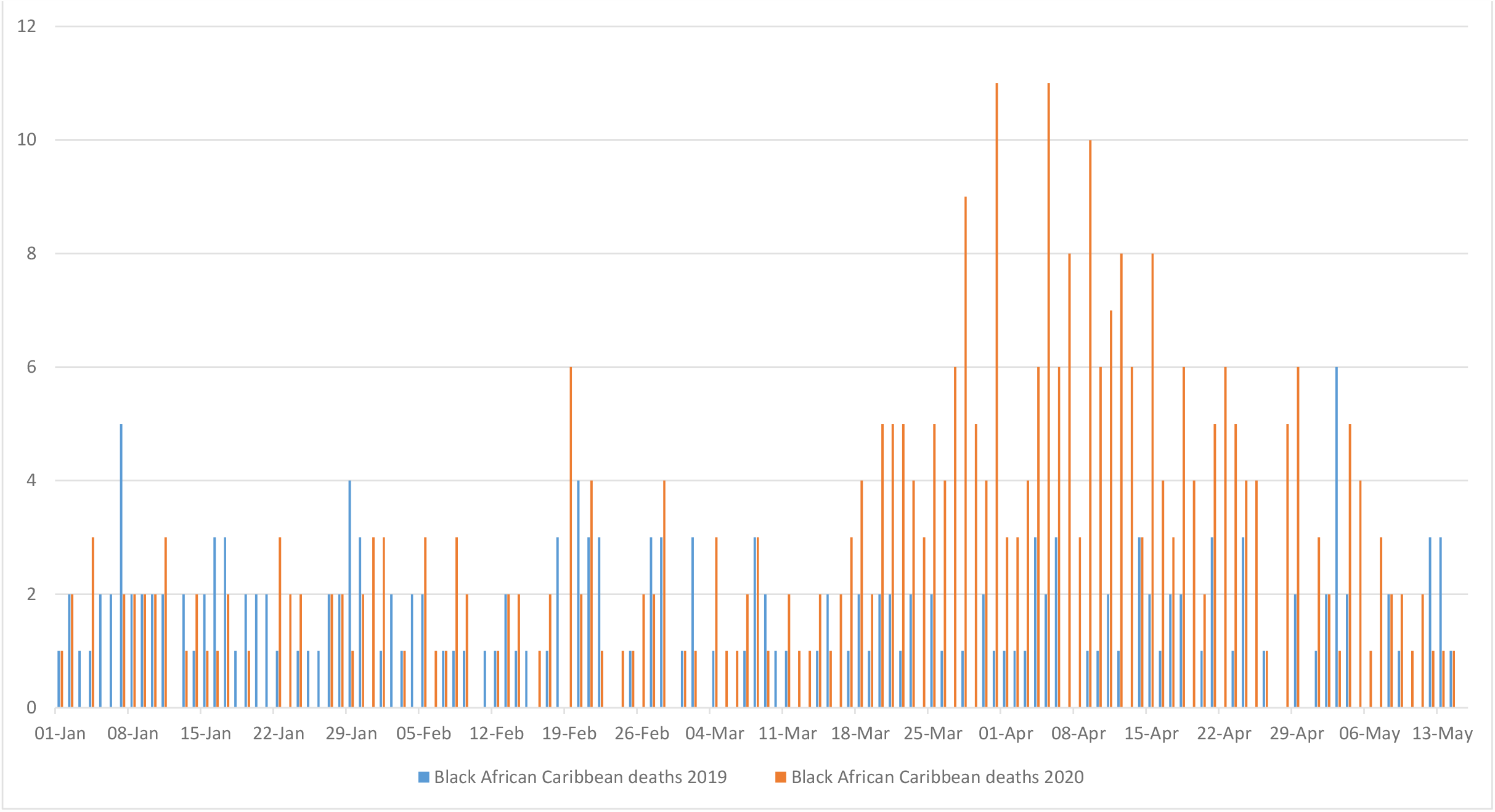
Daily mortality in mental health service users 2019 and 2020 -Black African/Caribbean.

**Figure 5:**
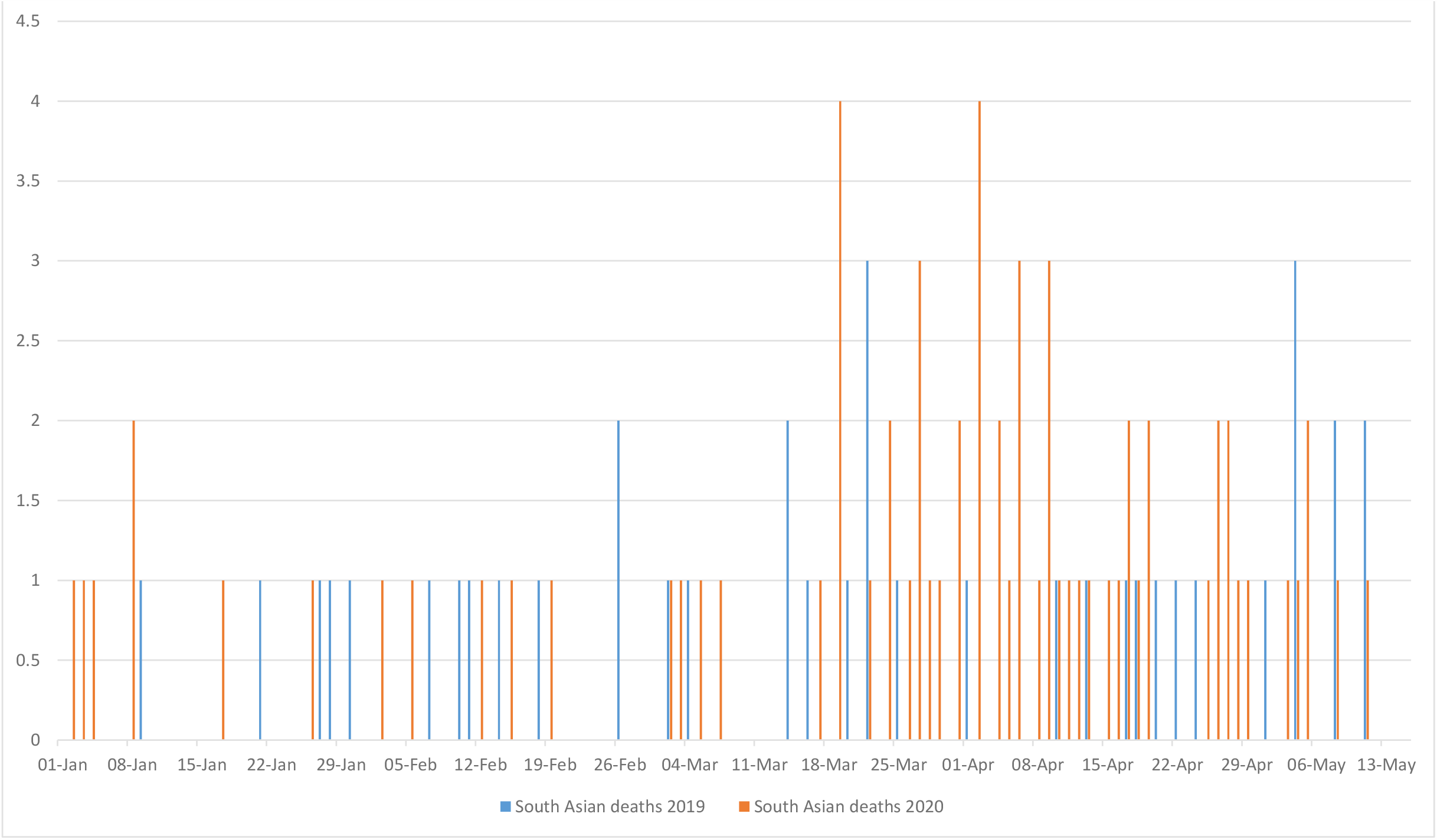
Daily mortality in mental health service users 2019 and 2020 -South Asian.

**Figure 6:**
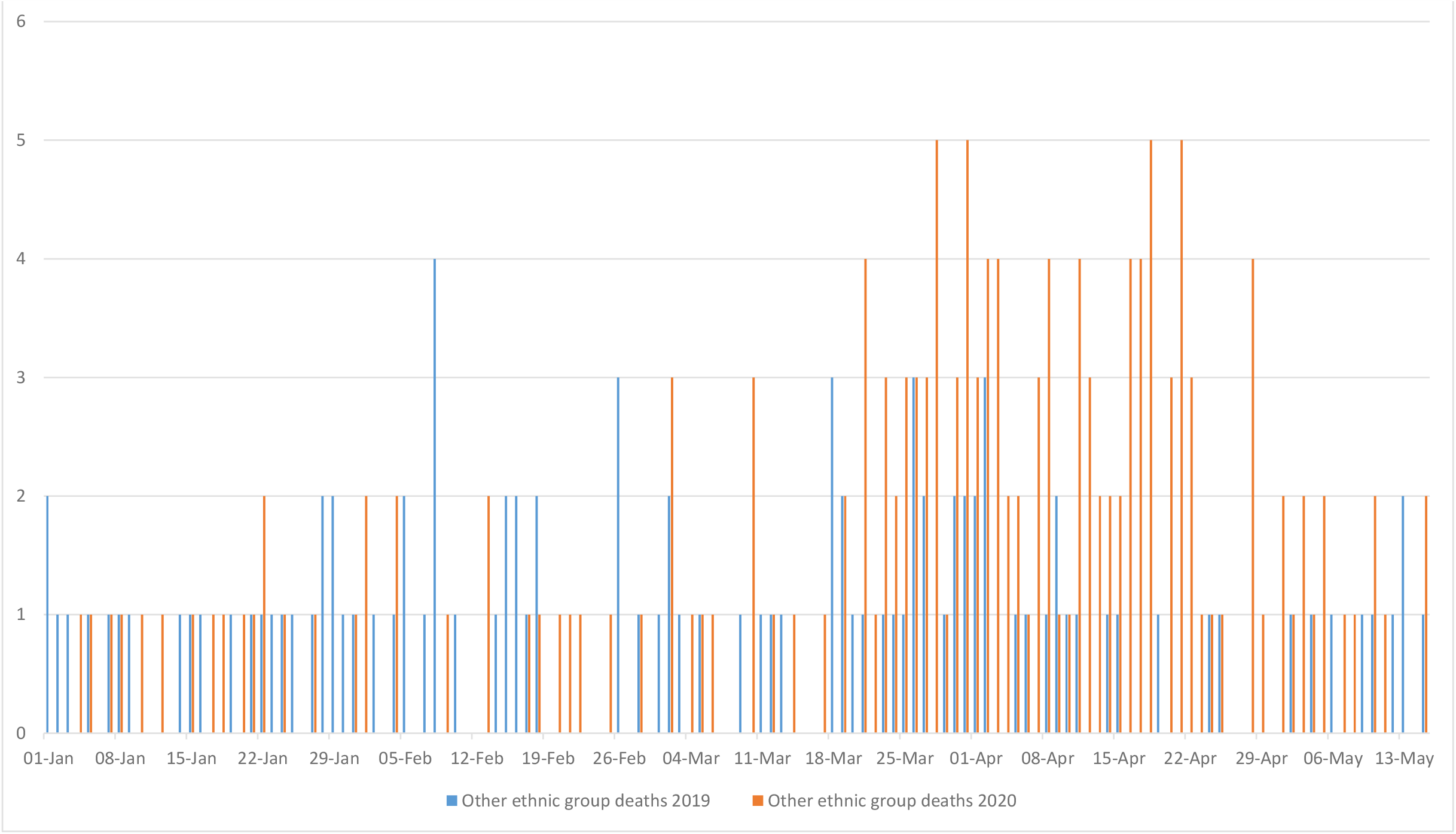
Daily mortality in mental health service users 2019 and 2020 -Other.

**Figure 7:**
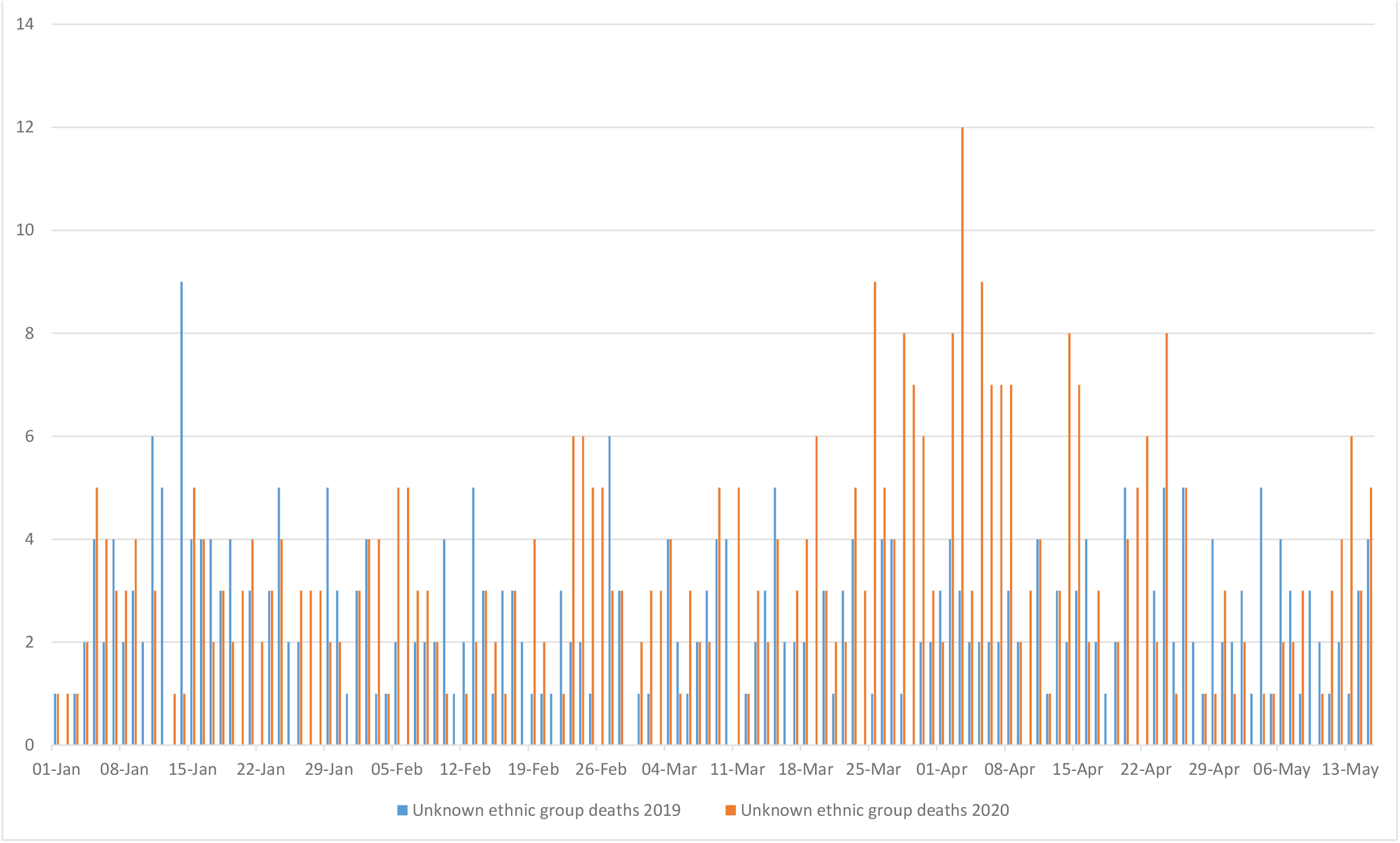
Daily mortality in mental health service users 2019 and 2020 -unknown ethnicity.

Distributions of ethnic groups in the different mortality groups are described in Table 1 and displayed graphically in Figure 8, showing an over-representation of Black African/Caribbean and Other ethnic groups in excess premature deaths compared to the total deaths.

**Figure 8:**
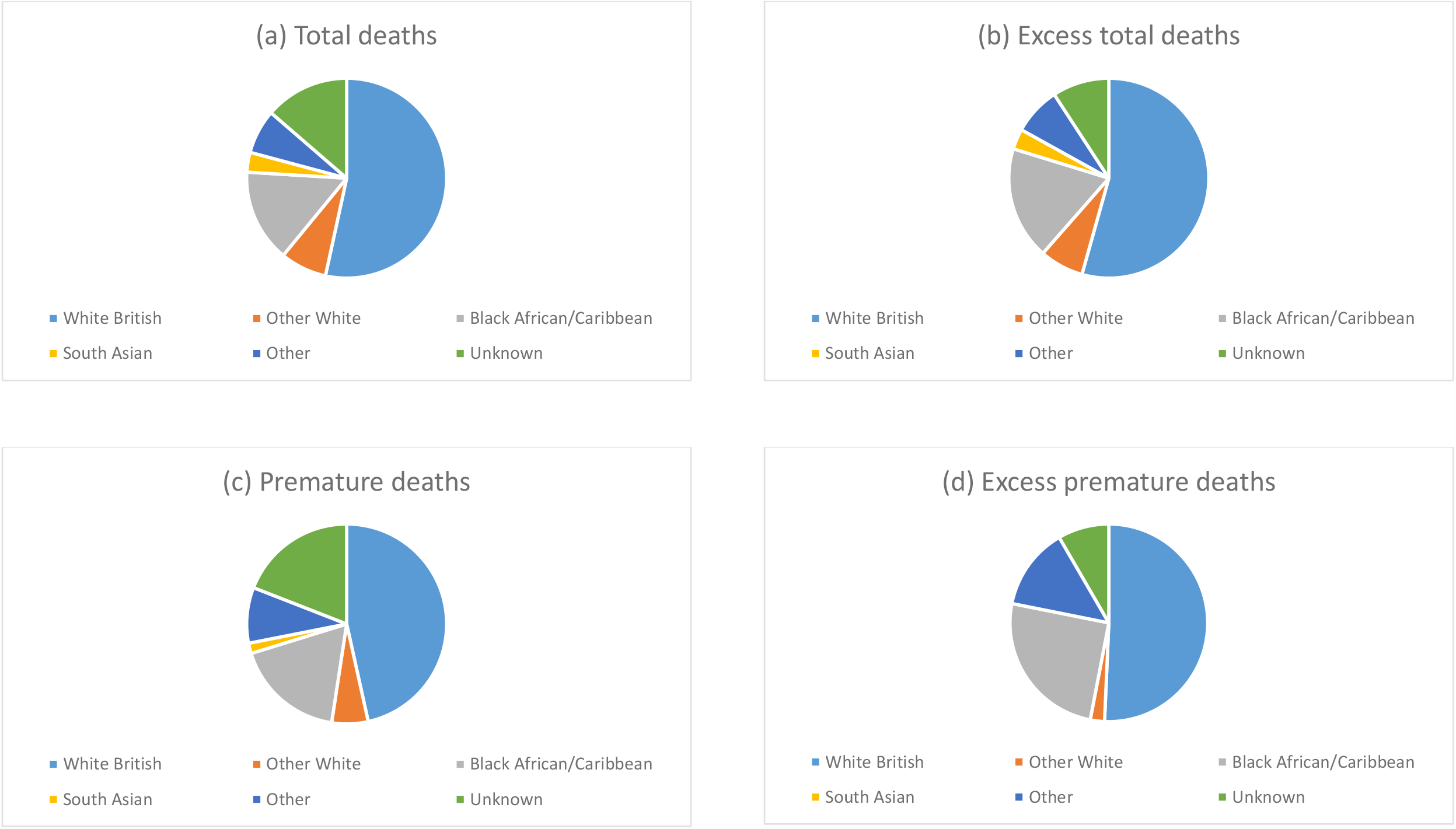
Ethnic group composition of mental health service users who died between 16.03.2020 and 15.05.2020.

## Discussion

Comparing the 2 months from 16^th^ March to 15^th^ May between 2020 and 2019, there was excess total and premature mortality in past and present SLaM service users. When examined by recorded ethnicity, the ratio of total deaths between the two years was highest at 3.33 in Black African/Caribbean patients and those from Other ethnic groups at 2.63, compared to 2.47 in White British patients. Considering premature deaths (where age at death was below 70 years), 2020:2019 ratios were 3.07 for Other ethnic groups and 2.74 for Black African/Caribbean patients compared to 1.96 for White British patients. Patients with recorded South Asian ethnicity had excess overall mortality in 2020 compared to 2019, with a similar ratio to that seen in White British patients, but lower numbers of premature deaths.

Considering limitations, it is important to bear in mind that the data are derived from a single site. Because complete data are being provided for that site with no hypothetical source population intended, calculation of confidence intervals was not felt to be appropriate for the descriptive data provided in this report; applicability to other mental healthcare providers cannot therefore be inferred and would need specific investigation. We additionally aim to provide further output on age-and sex-standardised mortality ratios in a future report. Profiles of services, patient populations and catchment morbidity are also likely to vary. Delays in national registrations might result in an underestimate of more recent deaths. No attempt was made to investigate modification or confounding by other factors such as age, gender, socioeconomic status or comorbid physical health conditions; however, these are unlikely to vary substantially between the at-risk population in 2019 and that in 2020. Mortality numbers clearly include all deaths and not just those attributable to COVID-19 infection. Finally, conflation of ethnic groups into the broad categories defined here was pragmatically driven by the need for sufficiently large group sizes. It is inevitably approximate and may fail to reflect varying experiences between constituent groups (e.g. between Black African and Black Caribbean groups; between Indian, Pakistani and Bangladeshi groups).

## Data Availability

Source data are available on request from the corresponding author.

## References

(1) Chang C-K, Hayes RD, Perera G, Broadbent MTM, Fernandes AC, Lee WE, et al. Life expectancy at birth for people with serious mental illness, substance use disorders, and depressive disorders from a secondary mental health care case register in London, UK. PLoS One 2011;6:e19590.

(2) Woodhead C, Ashworth M, Broadbent M, Callard F, Hotopf M, Schofield P, et al. Cardiovascular disease treatment among severe mental illness patients: a data linkage study between primary and secondary care. Br J Gen Practice 2016;66:e374–e381.

(3) Woodhead C, Cunningham R, Ashworth M, Barley E, Stewart R, Henderson M. Cervical and breast cancer screening uptake among women with serious mental illness: a data linkage study. BMC Cancer 2016;16:819.

(4) Holmes EA, O’Connor RC, Perry VH, Tracey I, Wessely S, Arseneault L, et al. Multidisciplinary research priorities for the COVID-19 pandemic: a call for action for mental health science. Lancet Psychiatry 2020;Apr 15:https://doi.org/10.1016/S2215-0366(20)30168-1.

(5) Stewart R, Martin E, Broadbent M. Mental health service activity during COVID-19 lockdown: South London and Maudsley data on working age community and home treatment team services and mortality from February to mid-May 2020. medRxiv 2020;doi: 10.1101/2020.06.13.20130419.

(6) Stewart R, Soremekun M, Perera G, Broadbent M, Callard F, Denis M, et al. The South London and Maudsley NHS Foundation Trust Biomedical Research Centre (SLAM BRC) Case Register: development and descriptive data. BMC Psychiatry 2009;9:51.

(7) Perera G, Broadbent M, Callard F, Chang C-K, Downs J, Dutta R, et al. Cohort profile of the South London and Maudsley NHS Foundation Trust Biomedical Research Centre (SLaM BRC) Case Register: current status and recent enhancement of an Electronic Mental Health Record derived data resource. BMJ Open 2016;6:e008721.

(8) Fernandes AC, Cloete D, Broadbent MTM, Hayes RD, Chang C-K, Roberts A, et al. Development and evaluation of a de-identification procedure for a case register sourced from mental health electronic records. BMC Med Inform Decis Mak 2013 Jan 1;13:71.

